# Association of age, sex, comorbidities, and clinical symptoms with the severity and mortality of COVID-19 cases: a meta-analysis with 85 studies and 67299 cases

**DOI:** 10.1101/2020.05.23.20110965

**Authors:** Mohammad Safiqul Islam, Md. Abdul Barek, Md. Abdul Aziz, Tutun Das Aka, Md. Jakaria

## Abstract

**Background:** A new pathogenic disease named COVID-19 became a global threat, first reported in Wuhan, China, in December 2019. The number of affected cases growing exponentially and now, more than 210 countries confirmed the cases.

**Objective:** This meta-analysis aims to evaluate risk factors, the prevalence of comorbidity, and clinical characteristics in COVID-19 death patients compared to survival patients that can be used as a reference for further research and clinical decisions.

**Methods:** PubMed, Science Direct, SAGE were searched to collect data about demographic, clinical characteristics, and comorbidities of confirmed COVID-19 patients from January 1, 2020, to May 17, 2020. Meta-analysis was performed with the use of Review Manager 5.3

**Results:** Eighty-five studies were included in Meta-analysis, including a total number of 67,299 patients with SARS-CoV-2 infection. Males are severely affected or died than females (OR = 2.26, p < 0.00001; OR = 3.59, p < 0.00001) are severely affected, or died by COVID-19 and cases with age ≥50 are at higher risk of death than age <50 years (OR=334.23). Presence of any comorbidity or comorbidities like hypertension, cardiovascular disease, diabetes, cerebrovascular disease, respiratory disease, kidney disease, liver disease, malignancy significantly increased the risk of death compared to survival (OR = 3.46, 3.16, 4.67, 2.45, 5.84, 2.68, 5.62, 2.81,2.16). Among the clinical characteristics such as fever, cough, myalgia, diarrhea, abdominal pain, dyspnea, fatigue, sputum production, chest tightness headache and nausea or vomiting, only fatigue (OR = 1.31, 95%) and dyspnea increased the death significantly (OR= 1.31, 4.57). The rate of death of COVID-19 cases is 0.03-times lower than the rate of survival (OR = 0.03).

**Conclusion:** Our result indicates that male patients are affected severely or died, the rate of death is more in the age ≥50 group, and the rate of death is affected by comorbidities and clinical symptoms.

## 1 Introduction

A cluster of critical cases with pneumonia arising from a new unknown pathogenic virus was first reported in Wuhan, Hubei, China, on December 31, 2019 [1]. Only a few days later, researchers have identified the virus which belongs to a relatively well-known viral family, Coronaviridae, and is similar to Bat SARS-like coronavirus (with 88% identity) and have identical nucleotide with the original SARS (80%), and MERS-Cov (about 50%) epidemic virus [2]. Based on phylogeny, taxonomy, and established practice, the International Committee on Taxonomy of Viruses named this pathogen as severe acute respiratory syndrome coronavirus-2 (SARS-CoV-2) [3]. Within a short time, novel coronavirus-infected pneumonia (NCIP) spread rapidly to the whole world. World Health Organization (WHO) declared NCIP as a Public and Global Health Emergency of Concern on January 30, 2020, and officially renamed this pandemic disease is coronavirus disease 2019 (COVID-19) on February 11, 2020 [3]. More than 210 countries already confirmed this case, and the case number is climbing rapidly day by day in the world reported by the WHO. As of May 17, 2020, about 4720196 cases with laboratory-confirmed COVID-19 infection have been detected, and 313220 (15%) died worldwide [4].

In China, the case number is gradually decreasing but rapidly increasing in other countries, especially in the USA, Italy, South Korea, and Iran. Coronavirus spike (S) glycoproteins bind with angiotensin-converting enzyme 2 (ACE2) receptors to promote entry into cells [5]. This new coronavirus is mainly transmitted through respiratory droplets and also can be transmitted by asymptomatic carriers. Several research studies report that this virus incubates 1 to 14 days in the human body [6]. It is essential to find out the clinical characteristics in the early stage to detect and isolate patients earlier and to minimize its transmission. Several clinical studies demonstrated that maximum confirmed patients had various symptoms (fever, cough, headache, dyspnea, myalgia, diarrhea, fatigue, nausea, and vomiting, etc.) with single or multiple comorbidities including cardiovascular disease, diabetes, hypertension, liver disease, kidney disease, and chronic obstructive pulmonary disease but asymptomatic individuals have also been identified as potential sources of infection [7]. The patients admitted to the intensive care unit (ICU) had a higher number of comorbidities than those not admitted to the ICU. SARS-CoV-2 can be transmitted through respiratory tract infections and develop severe pneumonia in infected patients like other viral respiratory infections. Severe patients may require intensive care and sometimes may result in death due to progressive respiratory failure. Everyone is susceptible to this virus, but the elderly and those with underlying diseases are more prone to adverse outcomes [8]. There are no approved drugs or vaccines in the world for fighting against COVID-19 [9]. Researchers are trying to develop medicine and vaccine to combat COVID-19 and upcoming SARS and MERS viruses [9]. There is an urgent need to find alternative ways of controlling the spread of this disease. Many researchers published clinical research on COVID-19, but the reported results were not all the same. However, for more accurate conclusions and further clinical practice, we collected the relevant studies about the clinical characteristics of COVID-19 to carry out an updated meta-analysis to describe epidemiological features, clinical characteristics, the prevalence of comorbidities, assess the risk of underlying diseases, and outcomes of death, as well as the survival of patients, confirmed to have the 2019-nCoV infection. Our findings may provide vital guidance for current clinical work on the prevention, treatment, and control of the current outbreak.

## 2 Methods

### 2.1 Search databases and search strategies

This meta-analysis was conducted by following the guidelines of PRISMA. The relevant studies were systematically searched in PubMed, ScienceDirect, and SAGE database from January 1, 2020, to May 17, 2020. EndNote X 7.0 software records are used to exclude duplicates. The following keywords are used in search alone or in combination: clinical characteristics of COVID-19, clinical outcome, death or clinical features, comorbidities of COVID-19, signs and symptoms of SARS-CoV-2. To identify the studies, there has no language or country limitation, and the search was restricted to humans, but only literature published online was included. We also checked reference lists of included studies to identify missing studies.

### 2.2 Study eligibility

#### 2.2.1 Inclusion criteria

1) Cohort studies and case series studies; 2) Study population confirmed with COVID-19; 3) The primary outcomes were: age, sex, clinical symptoms, signs, comorbidities, mortality; 4) No language and country restriction.

#### 2.2.2 Exclusion criteria

1) Reviews, editorials, letters, and commentaries; 2) Children and pregnant women case; 3) Overlapping or duplicate publications; 4) Irrelevant information for data extraction.

### 2.3 Data extraction and quality assessment

Rayyan QCRI, a systematic review web app, was used for the selection of the studies [10]. Literature search, evaluation, and data extraction to an excel database were conducted independently by two investigators (MAB and MAA) with the inclusion criteria. TDA and MSI resolved disagreements. Data extraction includes author, country, age, gender, number of participates, clinical symptoms, comorbidities, severity, death number, death patient’s age, gender, death number with comorbidities. Newcastle Ottawa Scale (NOS) Guidelines were followed for observational cohort studies to determine the quality of the included studies, as described elsewhere [11]. Jadad scale was used for randomized controlled trials (RCTs) to determine the consistency of the sample [12].

### 2.4 Statistical analysis

All analyses were performed by Microsoft Excel and Review Manager 5.3 (RevMan 5.3, The Cochrane Collaboration, Oxford, United Kingdom). Heterogeneity (χ^2^ and I^2^) between studies calculated using Review Manager 5.3. Heterogeneity in the forest plot was evaluated using both the Cochrane chi-square *Q*-test and I^2^ statistic. There was statistically significant heterogeneity if *p* < 0.1 or I^2^ > 50%. I^2^ values of 25%, 50%, and 75% represented low, moderate, and high heterogeneity, respectively. Sensitivity analysis was performed to quantify any significant variations in risk between the studies for each parameter by omitting studies one by one. Based on heterogeneity, the fixed effect model was used if I^2^ ≤ 50%, and the random effect model was used if I^2^>50%. Publication bias was evaluated by funnel plot along with Egger’s regression test. The level of significance of publication bias was p <0.05, the values larger than which were considered as no publication bias.

## 3. Results

### 3.1 Literature retrieval and Quality Assessment

Based on the initial search strategy, 2328 citations were found in different databases. A total of 539 records were retained after deleting duplicate records. Three hundred six articles were excluded after reviewing the title and abstract and 46 of the remaining 233 articles were excluded for various reasons. After a detailed assessment based on the inclusion criteria, finally, 85 articles [13-97] were included in the meta-analysis published in 2020, comprising of 67299 confirmed cases (Table 1). **Fig. 1** described the whole process of the selection of studies. It was also found that most of these studies (n=69) were based in China, although eight studies and six studies were identified from the USA and Italy, respectively. One study from South Korea and one study from Iran were also included. The quality of most included studies, assessed by the Newcastle Ottawa scale, was of high quality (score ranges between 6-8), with only two studies being of moderate quality (score 5), as shown in Table 2.

**Table 1.** Baseline characteristics of included studies.

| Study ID | Year | Country | Study Design | Sample Size | Male (%) | Median Age/<br>Mean±SD | Follow-up/Observation<br>period/Data<br>collection<br>period,<br>days |
| --- | --- | --- | --- | --- | --- | --- | --- |
| Aggarwal S | 2020 | USA | retrospective study | 7 | 12(75%) | 67/65.5 | 36 |
| Arentz M | 2020 | USA | retrospective study | 21 | 11 (52%) | 70 | 14 |
| Bellosta R | 2020 | Italy | single center,<br>observational<br>cohort study. | 20 | 18 (90%) | 75±9 | --- |
| Bezzio C | 2020 | Italy | prospective<br>observational<br>cohort study | 79 | 44 (557.7%) | 45 | 18 |
| Bhatraju PK | 2020 | USA | retrospective study | 24 | 15 (63%) | 64±18 | 13 |
| Cai Q | 2020 | China | retrospective study | 298 | 145 (48.66%) | 47.5 | 55 |
| Cai Y | 2020 | China | retrospective study | 7 | 5 (71.43%) | 60.29 | ----- |
| Cao J | 2020 | China | retrospective study | 102 | 53 (52%) | 54 | 30 |
| Chen T | 2020 | China | retrospective study | 274 | 171 (62%) | 62 | 30 |
| Chen G | 2020 | China | retrospective study | 21 | 17 (81%) | 56 | ----- |
| Chen T Dai Z | 2020 | China | a single-centered,<br>retrospective study | 203 | 108 (53.2%) | 54 | 41 |
| Chen X | 2020 | China | retrospective study | 48 | 37 (77.1%) | 64.6±18.1 | 19 |
| Chen Q | 2020 | China | single center<br>retrospective<br>observational study | 145 | 79 (54.5%) | 47.5 | 71 |
| Chu Y | 2020 | China | single center<br>retrospective<br>observational study | 33 | 22 (66.7%) | 65.2±16.6 | 29 |
| Colaneri M | 2020 | Italy | retrospective study | 44 | 28 (63.64%) | 67.5 | 13 |
| Deng Q | 2020 | China | retrospective study | 112 | 57 (50.9%) | 65.0 | 45 |
| Deng Y | 2020 | China | retrospective study | 225 | 124 (55.11%) | 54.5 | 52 |
| Duanmu Y | 2020 | USA | retrospective study | 100 | 56 (56%) | 45.0 | 19 |
| Du RH | 2020 | China | prospective cohort study | 179 | 97 (54.2) | 57.6±13.7 | 13 |
| Feng Y | 2020 | China | multi-center retrospective study | 476 | 271 (56.9%) | 53.0 | 46 |
| Goyal P | 2020 | USA | retrospective study | 393 | 238 (60.6%) | 62.2 | 24 |
| Grasselli G | 2020 | Italy | retrospective study | 1591 | 1304 (82%) | 63.0 | 34 |
| Guan WJ | 2020 | China | retrospective study | 1099 | 637 (57.96%) | 47.0 | 49 |
| Guo T | 2020 | China | single center retrospective study | 187 | 91 (48.7%) | 58.50 | 31 |
| He R | 2020 | China | retrospective study | 204 | 79 (38.73%) | 49.0 | 34 |
| Hong KS | 2020 | South Korea | Descriptive Study | 98 | 38 (38.8%) | 55.4±17.1 | 90 |
| Hu H | 2020 | China | retrospective study | 105 | 54 (50.94%) | 60.82±16.32 | 29 |
| Huang C | 2020 | China | prospective study | 41 | 30 (73%) | 49.0 | 17 |
| Jacobs JP | 2020 | USA | real time cohort study | 32 | 22 (68.8%) | 52.41 | 24 |
| Javanian M | 2020 | Iran | retrospective cohort study | 100 | 51 (51%) | 60.12 | 21 |
| Lei S | 2020 | China | retrospective study | 34 | 14 (41.2%) | 55.0 | 36 |
| Li X | 2020 | China | retrospective study | 548 | 279 (50.9%) | 60.0 | 37 |
| Li YK | 2020 | China | retrospective study | 25 | 13 (52%) | 61 | 51 |
| Liang W | 2020 | China | retrospective study | 1590 | 904 (57.4%) | 48.9 | 26 |
| Liu W | 2020 | China | retrospective study | 78 | 39 (50.0%) | 38.0 | 17 |
| Liu Y | 2020 | China | retrospective study | 109 | 59 (54.1%) | 55.0 | 41 |
| Liu Y Du X | 2020 | China | retrospective study | 245 | 114 (46.53%) | 53.95 | 60 |
| Liu F | 2020 | China | retrospective study | 140 | 49 (35.0%) | 65.5 | 54 |
| Liu J | 2020 | China | retrospective single-center study | 40 | 15 (37.5%) | 48.7±13.9 | 19 |
| Lodigiani C | 2020 | Italy | retrospective study | 388 | 264 (68%) | 66.0 | 57 |
| Lyu P | 2020 | China | retrospective study | 51 | 29 (56.86%) | 54±17 | 40 |
| Meng H | 2020 | China | retrospective study | 58 | 26 (44.8%) | 42.60±16.56 | 54 |
| Meng Y | 2020 | China | retrospective study | 168 | 86 (51.19%) | 56.7 | 19 |
| Mo P | 2020 | China | retrospective<br>single-center study | 155 | 86 (55.5%) | 54.0 | 36 |
| Niu S | 2020 | China | descriptive Study | 141 | 70 (49.65%) | 70.0 | 40 |
| Onder G | 2020 | Italy | ----- | 355 | 249 (70%) | 79.5 | ----- |
| Pan L | 2020 | China | a descriptive, cross-<br>sectional,<br>multicenter study | 204 | 107 (52.45%) | 52.9±16 | 60 |
| Peng S | 2020 | China | retrospective study | 11 | 8 (72.7%) | 61.0 | 24 |
| Peng YD | 2020 | China | retrospective study | 112 | 53 (47.32%) | 62.0 | 26 |
| Pereira MR | 2020 | USA | retrospective study | 90 | 53 (59%) | 57.0 | 20 |
| Richardson S | 2020 | USA | retrospective study | 5700 | 3437 (60.3%) | 63.0 | 35 |
| Shi S | 2020 | China | retrospective study | 671 | 322 (48.0%) | 63.0 | 54 |
| Shi H | 2020 | China | retrospective study | 81 | 42 (52%) | 49.5 | 33 |
| Shi Y | 2020 | China | retrospective study | 487 | 259 (53.2%) | 46.0 | 15 |
| Sun H | 2020 | China | retrospective study | 244 | 133 (54.51%) | 69.5 | 36 |
| Sun L | 2020 | China | retrospective study | 55 | 31 (56.4%) | 44.0 | 26 |
| Team NCPERE | 2020 | China | descriptive,<br>exploratory study | 44672 | 22,981<br>(51.4%) | ----- | 43 |
| Tian S | 2020 | China | retrospective study | 262 | 127 (48.5%) | 47.5 | 21 |
| Wan S | 2020 | China | retrospective study | 135 | 72 (53.3%) | 47 | 16 |
| Wang D | 2020 | China | retrospective<br>single-center study | 138 | 75 (54.3%) | 56 | 34 |
| Wang L | 2020 | China | retrospective<br>single-center study | 339 | 166 (48.97%) | 69.0 | 37 |
| Wang R | 2020 | China | single-center,<br>retrospective,<br>descriptive study | 125 | 54 (43.2%) | 38.76±13.80 | 29 |
| Wang X | 2020 | China | retrospective study | 1012 | 524 (51.8%) | 50.0 | 15 |
| Wang F | 2020 | China | retrospective study | 28 | 21 (75.0%) | 68.6±9.0 | 24 |
| Wang M | 2020 | China | retrospective study | 66 | 43 (65%) | 44.0 | 12 |
| Wu C | 2020 | China | retrospective study | 201 | 128 (63.7%) | 51.0 | 31 |
| Wu J | 2020 | China | retrospective study | 280 | 151 (53.93%) | 43.12 ±<br>19.02 | 31 |
| Xie H | 2020 | China | retrospective study | 79 | 44 (55.7%) | 60.0 | 21 |
| Xie J | 2020 | China | retrospective study | 56 | 24 (42.86%) | 56.5 | 10 |
| Xiong F | 2020 | China | retrospective study | 131 | 75 (57.3%) | 63.3 | 70 |
| Yan Y | 2020 | China | single-center,<br>retrospective,<br>observational study | 193 | 114 (59.1%) | 64.0 | 45 |
| Yang X | 2020 | China | a single-centered,<br>retrospective,<br>observational study | 52 | 35 (67%) | 59.7 | 28 |
| Yang Fa | 2020 | China | retrospective study | 52 | 28 (53.8%) | 63.0 | 106 |
| Yang Fb | 2020 | China | retrospective study | 92 | 49 (53.3%) | 69.8±14.5 | 50 |
| Yang Y | 2020 | China | retrospective study | 50 | 29 (58%) | 62.0 | 39 |
| Yao Q | 2020 | China | retrospective study | 108 | 43 (39.8%) | 52.0 | 12 |
| Yu X | 2020 | China | descriptive study | 333 | 172 (51.7%) | 56.0 | 26 |
| Yuan M | 2020 | China | descriptive study | 27 | 12 (45%) | 60.0 | 25 |
| Zhang JJ | 2020 | China | descriptive study | 140 | 71 (50.7%) | 57.0 | 18 |
| Zhang J | 2020 | China | retrospective<br>single-center study | 111 | 46 (41.44%) | 38.0 | 35 |
| Zheng F | 2020 | China | descriptive study | 161 | 80 (49.7%) | 45.0 | 21 |
| Zheng S | 2020 | China | retrospective study | 96 | 58 (60%) | 55.0 | 61 |
| Zhou F | 2020 | China | retrospective study | 191 | 119 (62%) | 56.0 | 34 |
| Zhou Y | 2020 | China | retrospective study | 21 | 13 (61.90%) | 66.10±13.94 | 34 |
| Zhao XY | 2020 | China | retrospective study | 91 | 49 (46.7%) | 46.0 | 25 |

**Figure 1.**
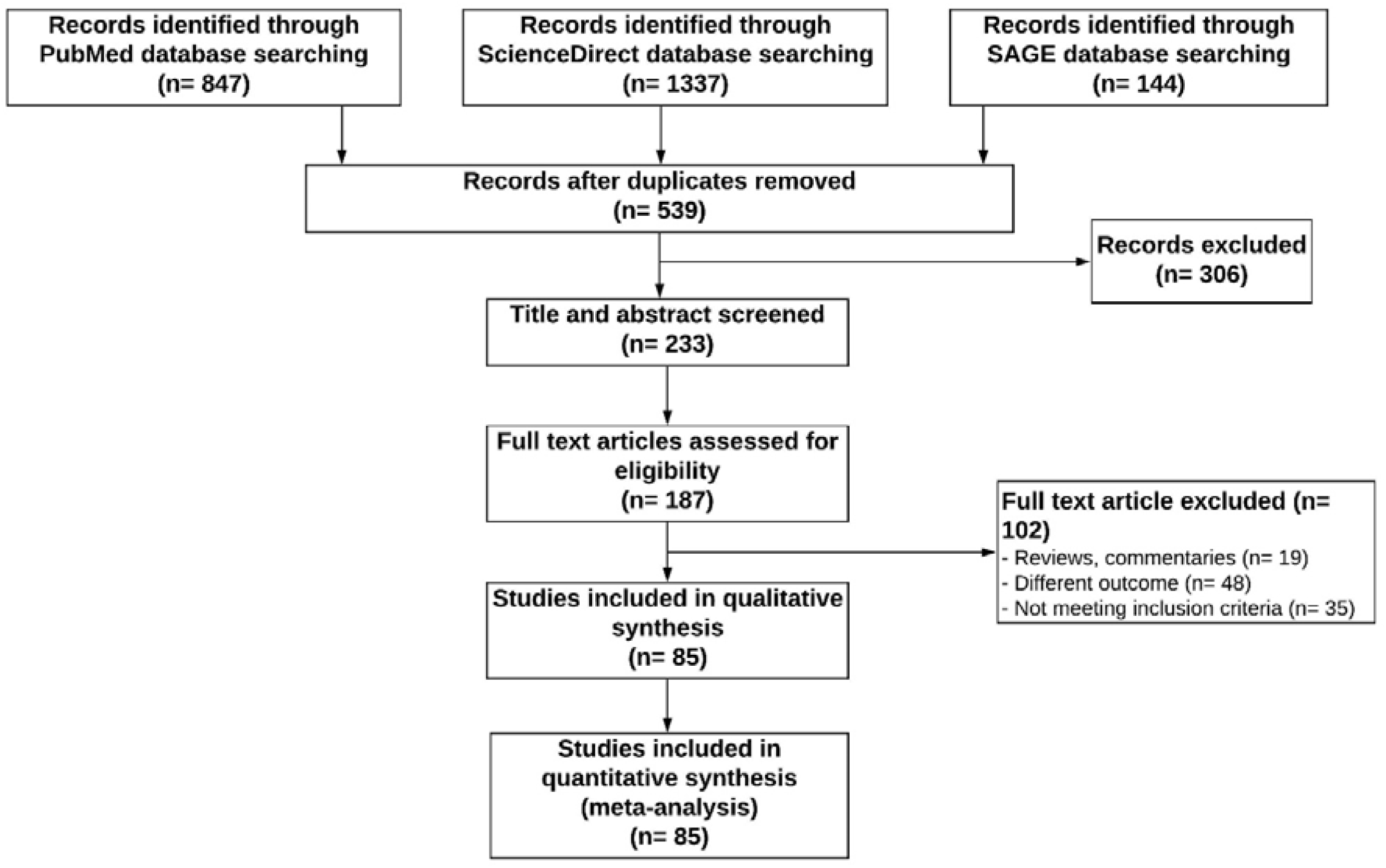
Flow diagram for selection of studies for inclusion in this meta-analysis

**Table 2.** Quality assessment of the included observational studies by NOS

| Author | Year | Selection |  |  |  | Comparability |  | Outcome |  |  | Total Score |
| --- | --- | --- | --- | --- | --- | --- | --- | --- | --- | --- | --- |
|  |  | 1 | 2 | 3 | 4 | 5A | 5B | 6 | 7 | 8 |  |
|  |  | Exposed cohort truly/somewhat representative | Nonexposed cohort drawn from the same community | Ascertainment of exposure | Outcome of interest not present at start | Cohorts adjusted for age | Cohorts adjusted for other important factor(s) | Quality of outcome assessment | Follow-up/ Observation period Long enough for outcomes to occur | Adequacy of follow-up of cohorts |  |
| Aggarwal S | 2020 | * | * | * | * | * |  | * | * | - | 7 |
| Arentz M | 2020 | * | * | * | * |  |  | * | * | - | 6 |
| Bellosta R | 2020 | * | - | * | * | * |  | * | * | - | 6 |
| Bezzio C | 2020 | * | * | * | * | * | * | * | * | - | 8 |
| Bhatraju PK | 2020 | * | - | * | * | * |  | * | * | - | 6 |
| Cai Q | 2020 | * | - | * | * |  | * | * | * | * | 7 |
| Cai Y | 2020 | * | - | * | * | * | * | * | * | * | 8 |
| Cao J | 2020 | * | - | * | * |  |  | * | * | * | 6 |
| Chen T | 2020 | * |  | * | * |  |  | * | * | * | 6 |
| Chen G | 2020 | * | * |  | * |  |  | * | * | * | 7 |
| Chen T Dai Z | 2020 | * |  | * | * |  |  | * | * | * | 6 |
| Chen X | 2020 | * | * |  | * | * |  | * | * |  | 6 |
| Chen Q | 2020 | * | * | * | * |  |  | * | * |  | 6 |
| Chu Y | 2020 | * | * | * | * |  |  | * | * | * | 7 |
| Colaneri M | 2020 | * |  | * | * |  | * | * | * | * | 7 |
| Deng Q | 2020 | * |  | * | * |  |  | * | * | * | 6 |
| Deng Y | 2020 | * | * | * | * |  | * | * | * | * | 8 |
| Duanmu Y | 2020 | * | * | * | * |  |  | * | * |  | 6 |
| Du RH | 2020 | * | * | * | * |  |  | * | * |  | 6 |
| Feng Y | 2020 | * |  | * | * |  |  | * | * | * | 6 |
| Goyal P | 2020 | * |  | * | * | * |  | * | * |  | 6 |
| Grasselli G | 2020 | * | * | * | * |  |  | * | * |  | 6 |
| Guan WJ | 2020 | * | * | * | * |  |  | * | * |  | 6 |
| Guo T | 2020 | * |  | * | * |  |  | * | * | * | 6 |
| He R | 2020 | * | * | * | * |  |  | * | * |  | 6 |
| Hong KS | 2020 | * |  | * | * |  |  | * | * |  | 5 |
| Hu H | 2020 | * | * | * | * |  |  | * | * |  | 6 |
| Huang C | 2020 | * | * | * | * |  |  | * | * |  | 6 |
| Jacobs JP | 2020 | * | * | * | * |  | * | * | * |  | 7 |
| Javanian M | 2020 | * | * | * | * |  |  | * | * |  | 6 |
| Lei S | 2020 | * |  | * | * |  | * | * | * |  | 6 |
| Li X | 2020 | * | * | * | * |  |  | * | * |  | 6 |
| Li YK | 2020 | * |  | * | * |  |  | * | * |  | 6 |
| Liang W | 2020 | * |  | * | * | * |  | * | * |  | 6 |
| Liu W | 2020 | * |  | * | * | * | * | * | * |  | 7 |
| Liu Y | 2020 | * |  | * | * | * |  | * | * |  | 6 |
| Liu Y Du X | 2020 | * | * | * | * |  |  | * | * |  | 6 |
| Liu F | 2020 | * |  | * | * | * |  | * | * |  | 6 |
| Liu J | 2020 | * | * | * | * |  | * | * | * |  | 7 |
| Lodigiani C | 2020 | * |  | * | * | * |  | * | * |  | 6 |
| Lyu P | 2020 | * |  | * | * | * | * | * | * |  | 7 |
| Meng H | 2020 | * |  | * | * |  |  | * | * | * | 6 |
| Meng Y | 2020 | * |  | * | * |  |  | * | * |  | 6 |
| Mo P | 2020 | * | * | * | * |  |  | * | * |  | 6 |
| Niu S | 2020 | * |  | * | * |  |  | * | * |  | 6 |
| Onder G | 2020 | * |  | * | * | * |  | * | * |  | 6 |
| Pan L | 2020 | * |  | * | * |  |  | * | * |  | 6 |
| Peng S | 2020 | * | * | * | * |  | * | * | * |  | 7 |
| Peng YD | 2020 | * |  | * | * |  | * | * | * |  | 6 |
| Pereira MR | 2020 | * |  | * | * | * |  | * | * |  | 6 |
| Richardson S | 2020 | * |  | * | * | * |  | * | * |  | 6 |
| Shi S | 2020 | * |  | * | * | * | * | * | * |  | 7 |
| Shi H | 2020 | * |  | * | * |  |  | * | * | * | 6 |
| Shi Y | 2020 | * |  | * | * | * |  | * | * |  | 6 |
| Sun H | 2020 | * | * | * | * | * |  | * | * |  | 7 |
| Sun L | 2020 | * |  | * | * |  | * | * | * |  | 6 |
| Team NCPERE | 2020 | * |  | * | * |  |  | * | * | * | 6 |
| Tian S | 2020 | * |  | * | * |  |  | * | * | * | 6 |
| Wan S | 2020 | * | * | * | * |  |  | * | * |  | 6 |
| Wang D | 2020 | * |  | * | * |  | * | * | * | * | 7 |
| Wang L | 2020 | * | * | * | * |  |  | * | * |  | 6 |
| Wang R | 2020 | * |  | * | * | * |  | * | * | * | 7 |
| Wang X | 2020 | * |  | * | * |  |  | * | * | * | 6 |
| Wang F | 2020 | * |  | * | * |  |  | * | * | * | 6 |
| Wang M | 2020 | * |  | * | * |  | * | * | * |  | 6 |
| Wu C | 2020 | * |  | * | * | * | * | * | * | * | 8 |
| Wu J | 2020 | * | * | * | * |  |  | * | * |  | 6 |
| Xie H | 2020 | * |  | * | * |  |  | * | * | * | 6 |
| Xie J | 2020 | * |  | * | * |  |  | * | * |  | 6 |
| Xiong F | 2020 | * |  | * | * | * |  | * | * |  | 6 |
| Yan Y | 2020 | * | * | * | * |  |  | * | * |  | 7 |
| Yang X | 2020 | * |  | * | * |  | * | * | * |  | 6 |
| Yang Fa | 2020 | * |  | * | * |  |  | * | * | * | 6 |
| Yang Fb | 2020 | * | * | * | * |  |  | * | * | * | 7 |
| Yang Y | 2020 | * | * | * | * |  |  | * | * |  | 6 |
| Yao Q | 2020 | * | * | * | * |  |  | * | * |  | 6 |
| Yu X | 2020 | * |  | * | * | * |  | * | * |  | 6 |
| Yuan M | 2020 | * |  | * | * | * |  | * | * |  | 6 |
| Zhang JJ | 2020 | * | * | * | * |  |  | * | * |  | 6 |
| Zhang J | 2020 | * |  | * | * |  |  | * | * | * | 6 |
| Zhao XY | 2020 | * |  | * | * |  |  | * | * |  | 5 |
| Zheng F | 2020 | * | * | * | * | * | * | * | * |  | 8 |
| Zheng S | 2020 | * |  | * | * | * |  | * | * |  | 6 |
| Zhou F | 2020 | * | * | * | * |  |  | * | * |  | 6 |
| Zhou Y | 2020 | * | * | * | * |  |  | * | * |  | 6 |

NOS=Newcastle Ottawa Scale; in this scale score ranges between 0-9 where “0-3” indicates low quality, “4-5” indicates moderate quality and score of $\geq 6$ indicates high quality study

### 3.2 Effect of sex on disease severity

Fifty-five studies reported the sex wise disease severity, and significant heterogeneity was found when the severity of the male and female COVID-19 patients were compared (I^2^ = 79%, p<0.00001). The random-effect model was used in the meta-analysis and the proportion of severe patients in the male group was significantly higher than the female group, and male patients showed 2.26-times more risk of death than female patients (male vs. female; 57.74 vs. 42.36%, OR = 2.26; 95% CI = 1.79–2.86; p<0.00001) (**Fig. 2**).

**Figure 2.**
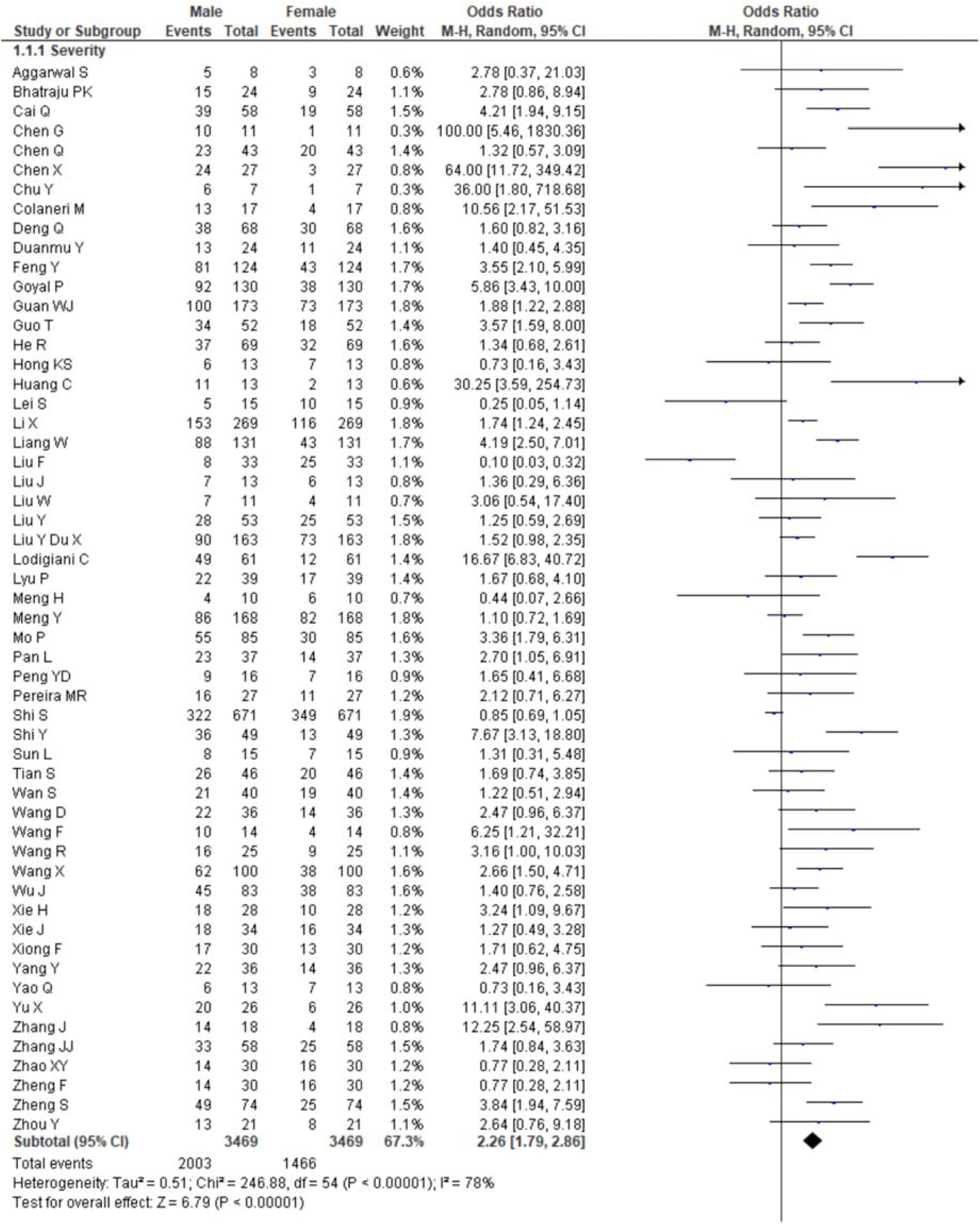
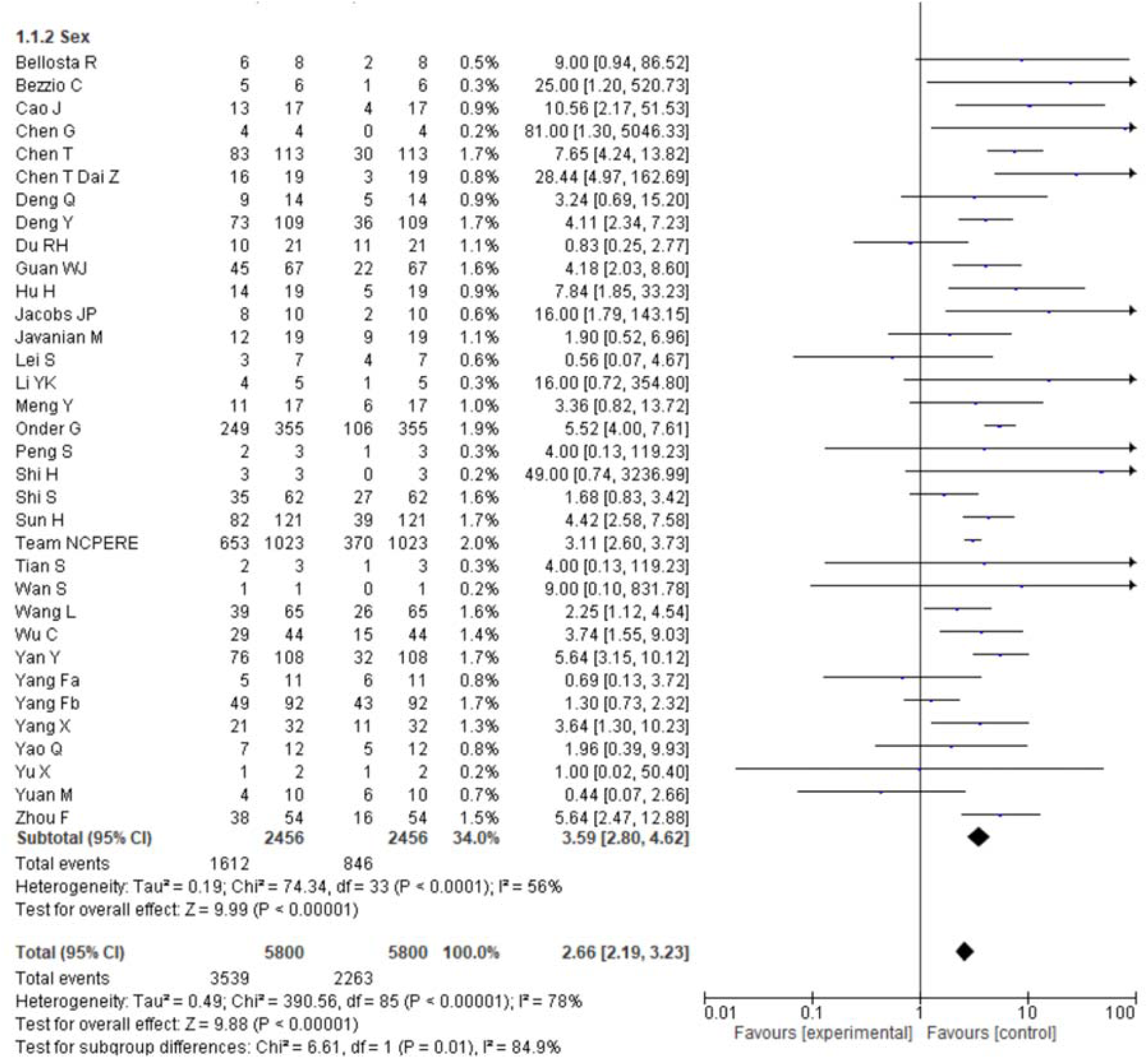
Pooled effects of sex on COVID-19-linked death. All the studies reported death as a clinical outcome except the Guan G where the outcome was recorded as a primary composite endpoint consisting of death, admission to an intensive care unit or use of mechanical ventilation.

### 3.3 Effect of sex on death

A total of thirty-four studies described the sex wise death report. Male patients with COVID-19 shown 3.59-times more risk of death compared to female patients (male vs. female; 65.45 vs. 34.45%, OR = 3.59; 95% CI = 2.80-4.62, p<0.00001). Significant heterogeneity was also reported in this finding (I^2^ = 56%, p<0.0001). (**Fig. 2**).

### 3.4 Effect of age on the death

Among the 85 studies, 41 studies reported death according to the age range, and only 152 deaths in females were found within a total of 3405 deaths. A higher significant heterogeneity also found in this case (I^2^ = 87%, p<0.0001). Covid-19 patients age more than 50 years showed 334.23 times more risk of death in comparison with COVID-19 patients aged below 50 years (age ≥50 years vs. age <50 years: 95.54 vs. 4.46%, OR = 324.23; 95% CI = 139.25-802.20, p<0.00001). (**Fig. 3**)

**Figure 3.**
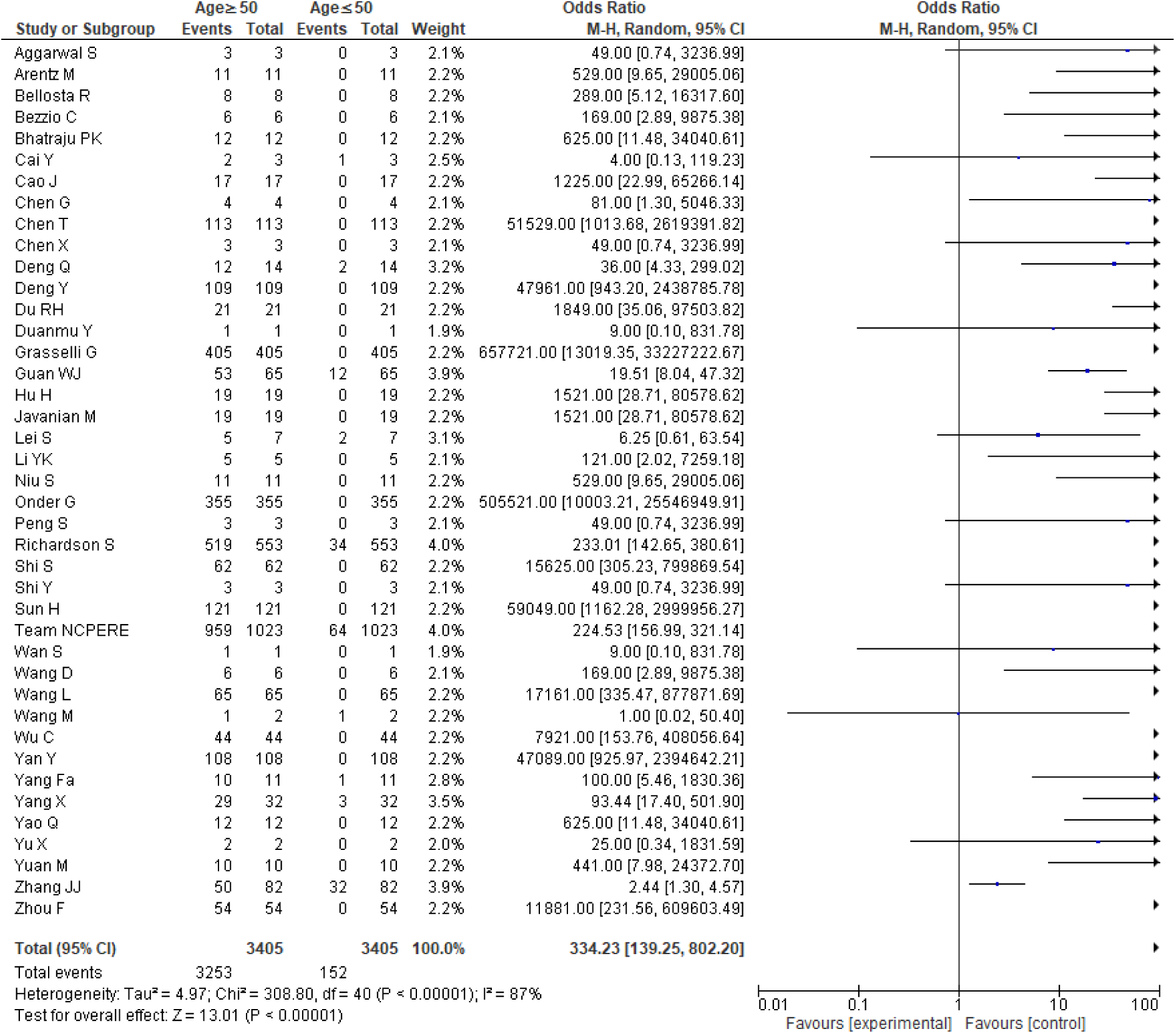
Pooled effects of age on COVID-19-linked death (age ≥50 Vs. age<50 years)

### 3.5 Comorbidity

The prevalence of comorbidities including hypertension, diabetes, respiratory disease, cardiovascular diseases, cerebrovascular disease, renal disease, liver disease, malignancy, and gastrointestinal (GI) disease in death and survived patients of the included studies is shown in **Fig. 4**. Among the different comorbidities, 68.25% of patient death was due to the presence of at least one comorbidity, and patients having at least one comorbidity had 3.46-times risk of death than survived patients (OR = 3.46, 95% CI =2.56-4.67, p < 0.00001).

**Figure 4.**
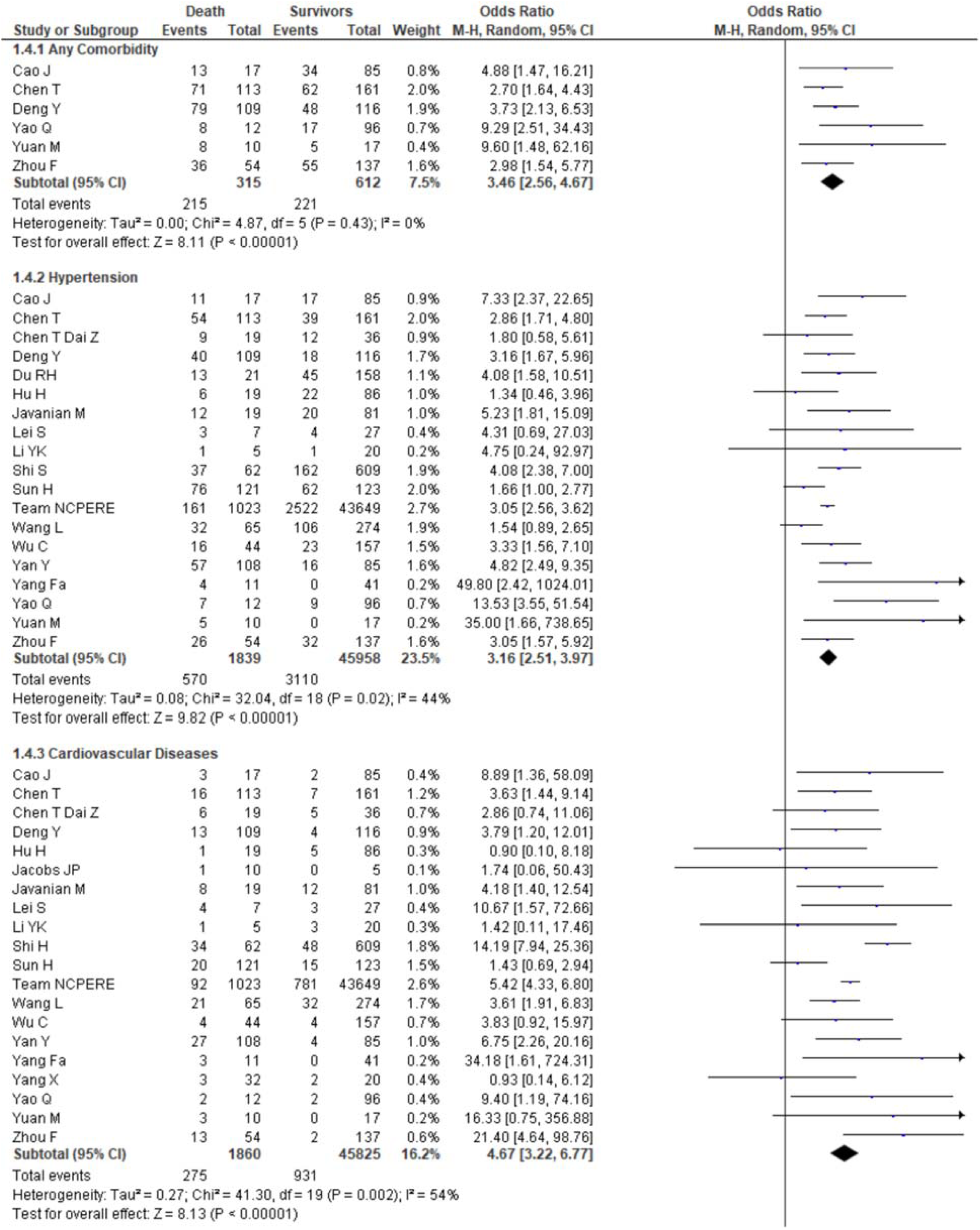
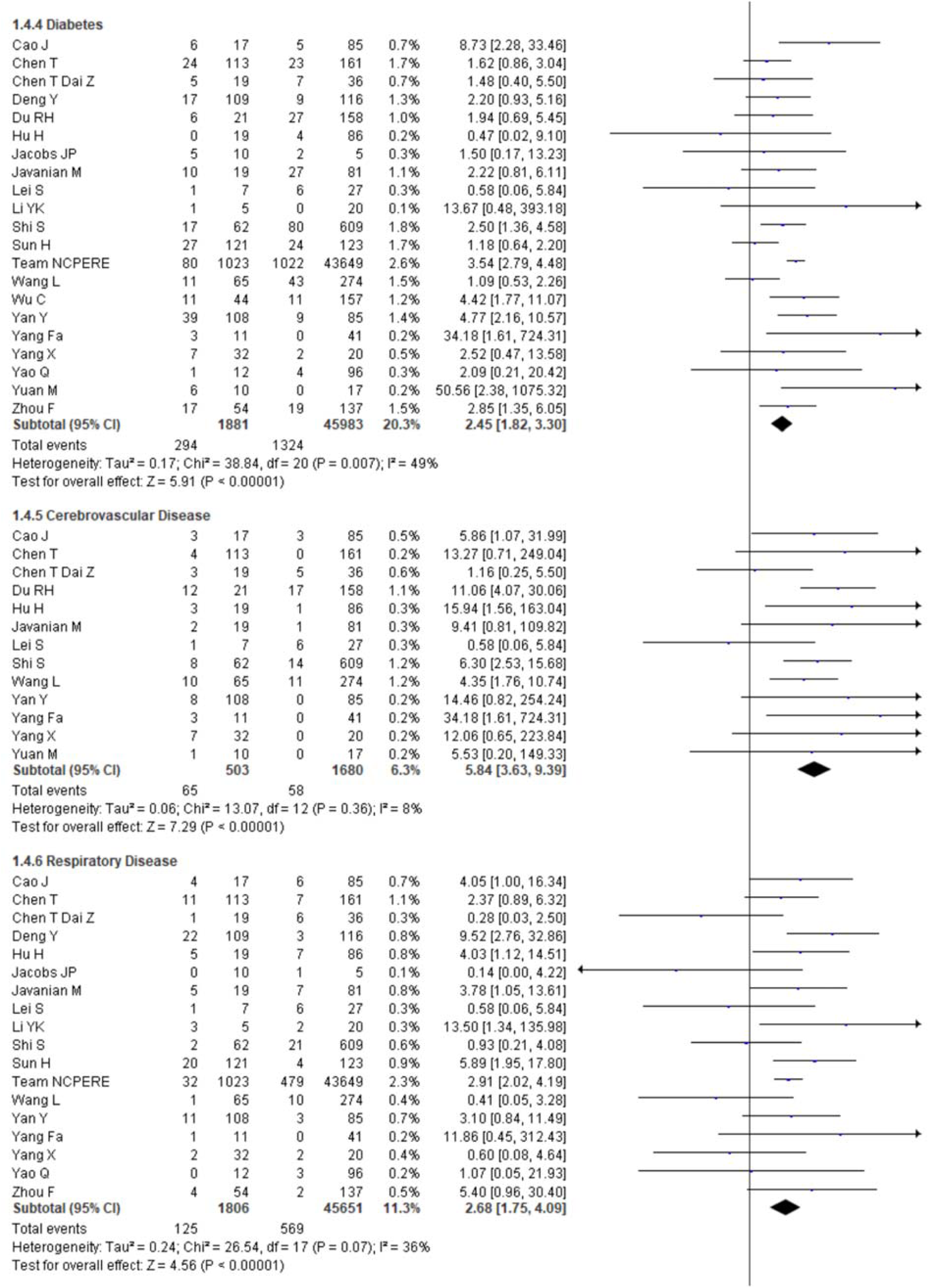
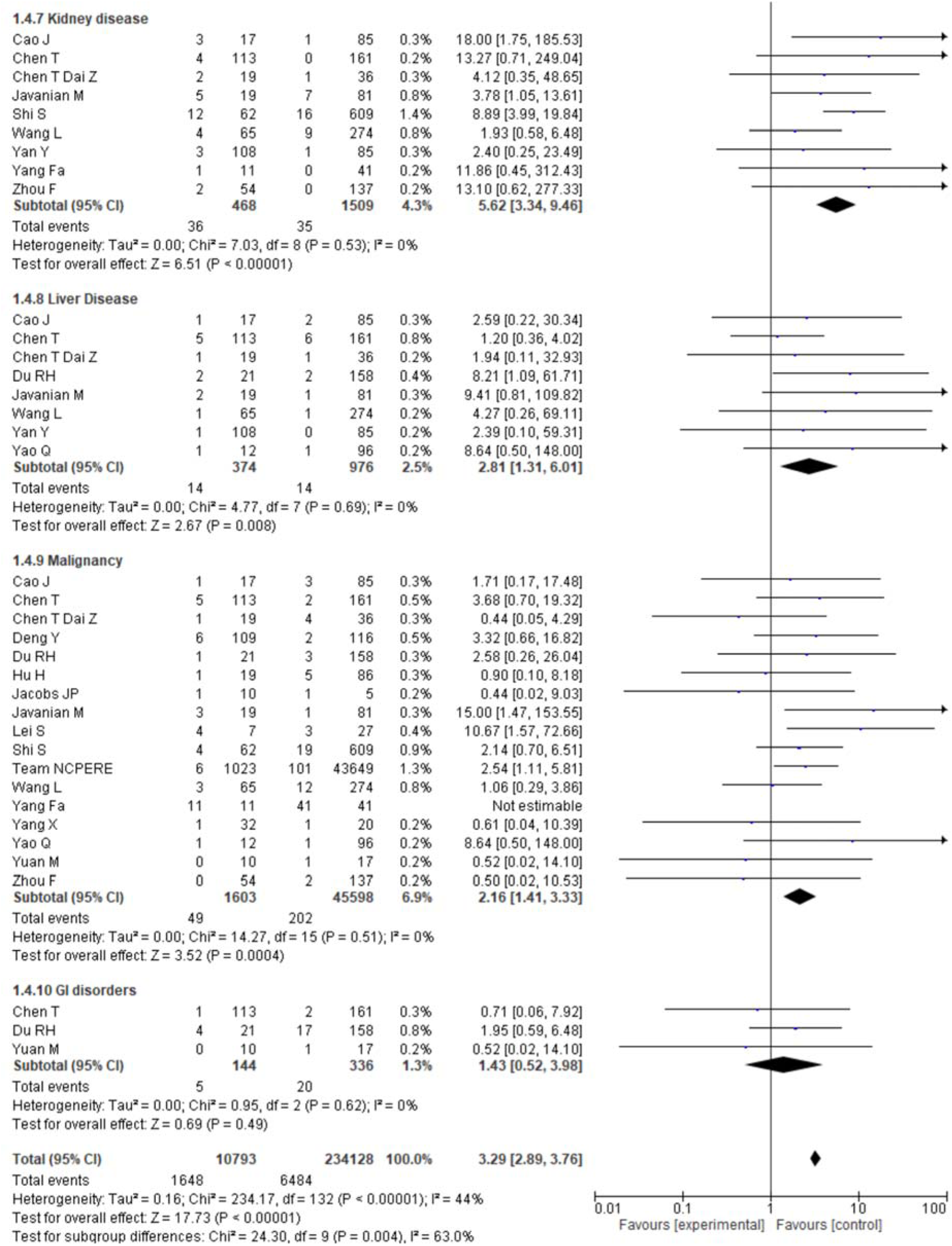
Pooled effects of comorbidities on the death linked with COVID-19

A total of 31% death was found due to hypertension in comparison to the survival (6.77%) with hypertension, and the chance of death was found 3.16-times higher in comparison to survival patients with hypertension (OR = 3.16, 95% CI = 2.51-3.97, p < 0.00001). The proportion of death in cerebrovascular disease and cardiovascular disease was also higher than the survival rate with these diseases (12.92% vs. 14.78% death, OR = 5.84, 95% CI = 3.63-9.39), p< 0.00001; OR = 4.67, 95% CI = 3.22-6.77, p < 0.00001, respectively). The percentages of death with diabetes, respiratory disease, kidney disease, and liver diseases were significantly higher than the survival rates with these diseases (death vs. survival: 15.63 vs. 2.66%; 6.92 vs. 1.25%; 7.69 vs. 2.32%; 3.74 vs. 1.43%, respectively). Covid-19 patients with diabetes, respiratory disease, kidney disease, and liver diseases had significantly 2.45, 2.68, 5.62 and 2.81-times more risk of death, respectively, compared to survival with these diseases.

The percentage of survival of COVID-19 patients with malignancy is very low (0.44%) and COVID-19 patients with malignancy had 2.16-times more risk of death compared to survival (OR= 2.16, 95% Cl = 1.41-3.33, p=0.0004).

No association was found with the death of COVID-19 patients having GI disease as comorbidity (p>0.05). Overall, the Covid-19 patients with comorbidity had 3.29-times more significant risk of death than survival (OR = 3.29, 95% Cl = 2.89-3.76, p <0.00001).

### 3.6 Clinical Manifestation

Fourteen studies reported each of fever and cough, 12 studies reported fatigue, 11 studies reported each of dyspnea and diarrhea, 8 studies reported each of myalgia and headache, 6 studies reported nausea or vomiting, 5 studies reported each of sputum production and chest tightness and 4 studies reported abdominal pain. The most prevalent clinical symptoms were fever (91.68%) and cough (74.4%) followed by dyspnea (47.71%), fatigue (45.94%), sputum production (37.56%), chest tightness (36.90%), myalgia (23.73%), diarrhea (22.44%), headache (14.73%), nausea or vomiting (13.28%), and abdominal pain (12.29%). Among the different clinical symptoms, fatigue (OR = 1.31, 95% Cl = 1.04-1.66, p = 0.02) and dyspnea increased the death significantly (OR = 4.57, 95% Cl = 2.69-7.78, p <0.0001). The chest tightness remained in the borderline (OR = 2.04, 95% Cl = 0.994.20, p = 0.05), whereas no association of death was found with the other clinical symptoms (**Fig. 5**). The overall impact of clinical symptoms on death is also high (OR = 1.25, 95%Cl = 1.06-1.48, p = 0.007) with a significant heterogeneity (p <0.00001).

**Figure 5.**
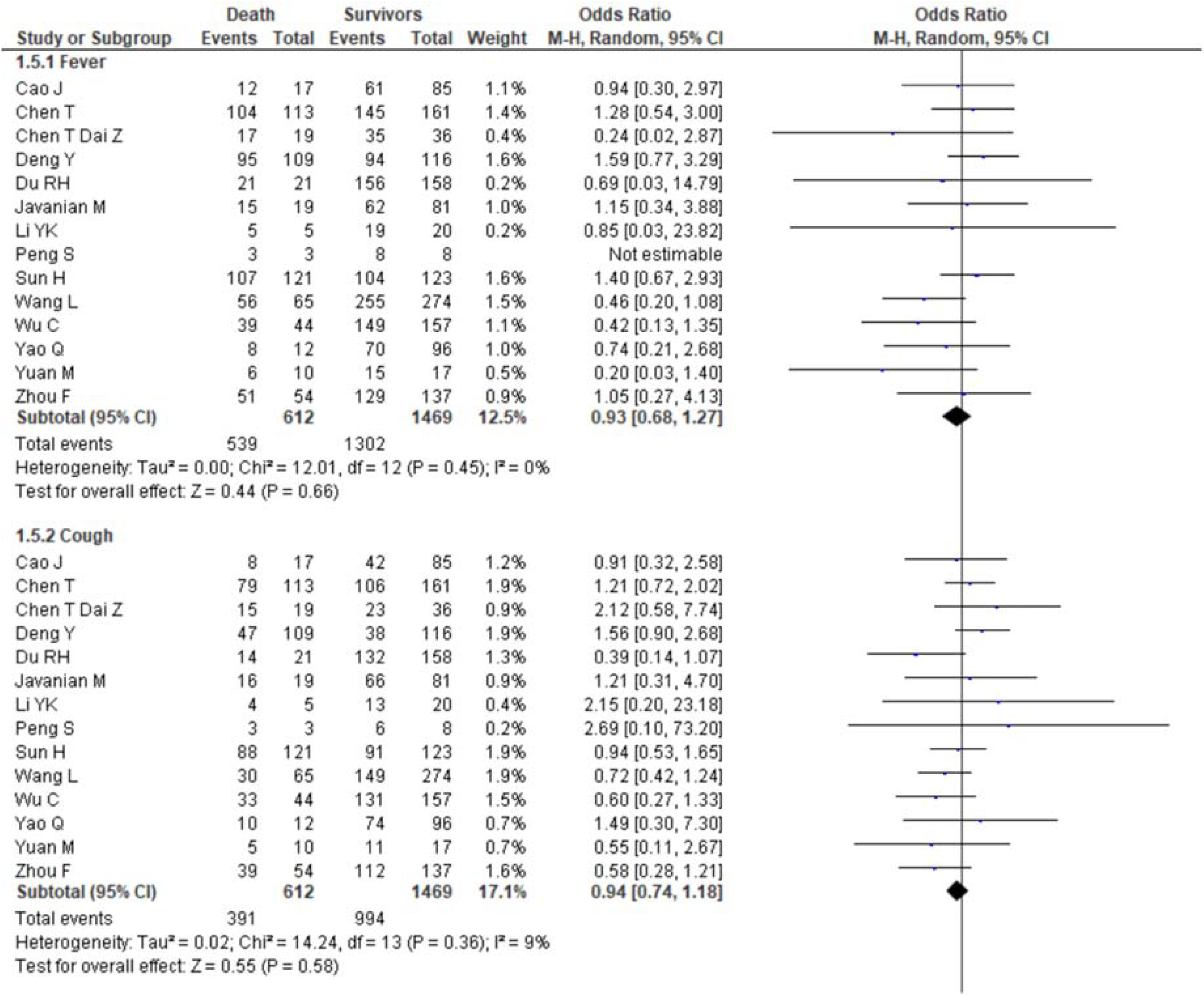
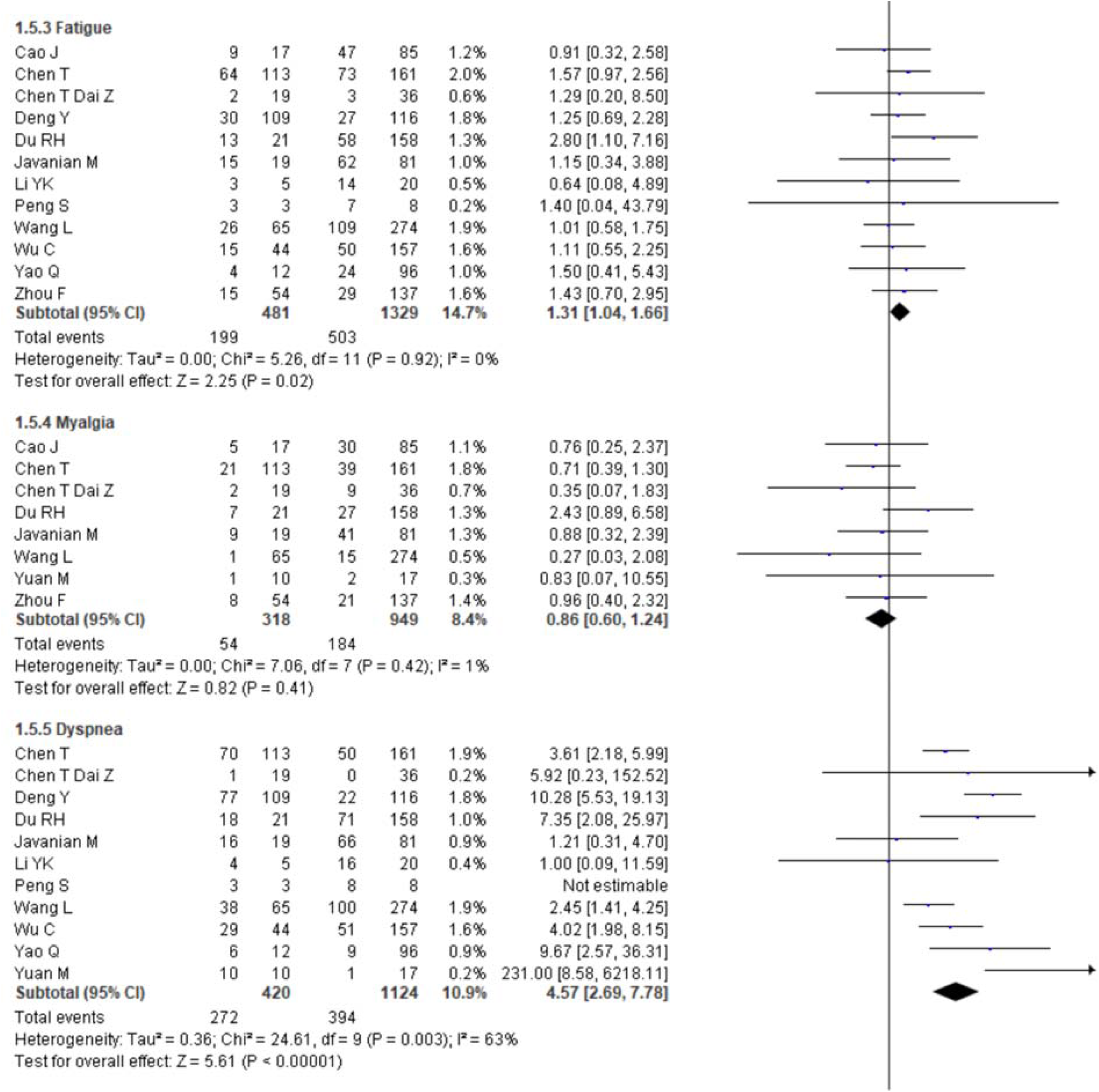
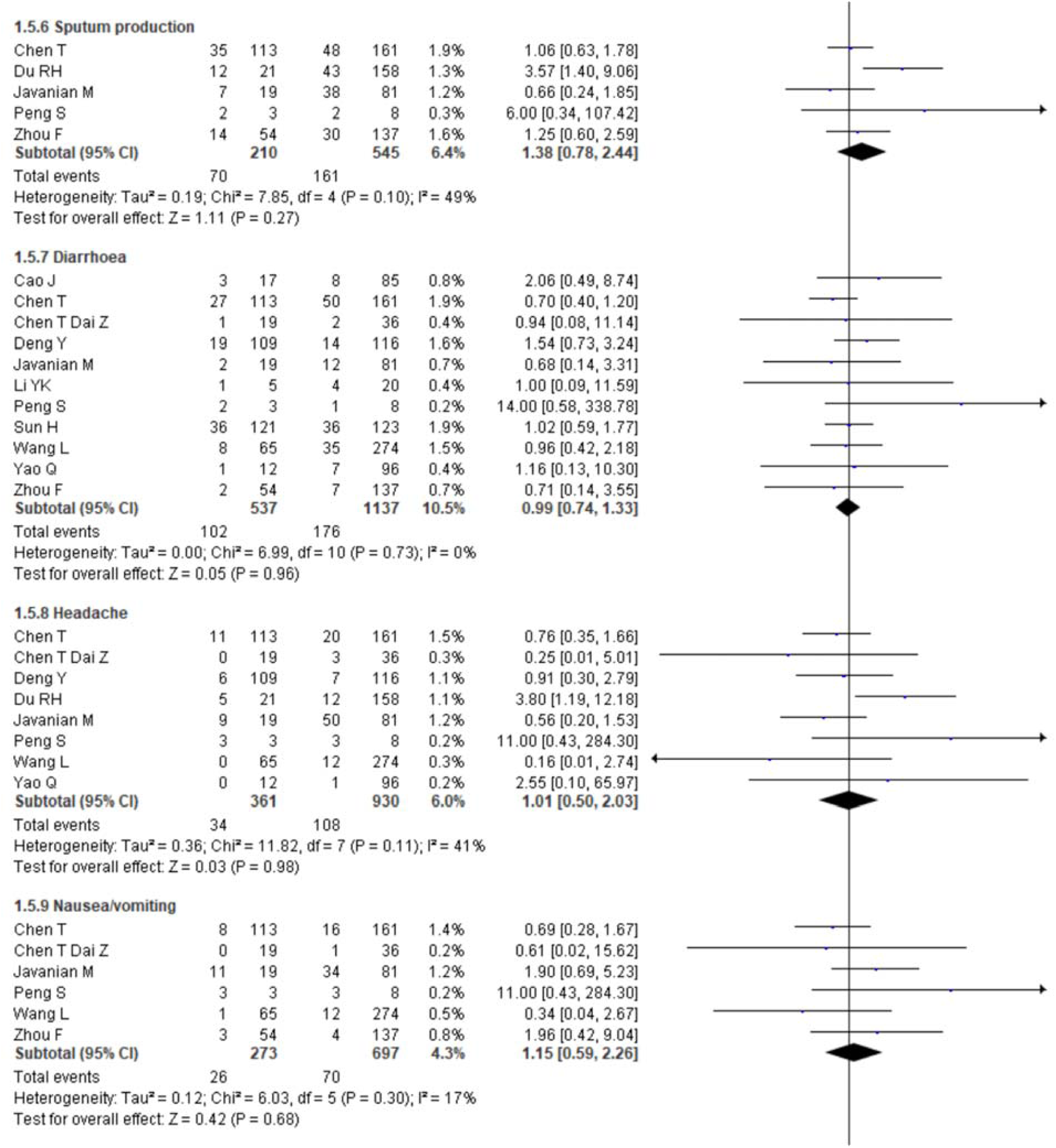
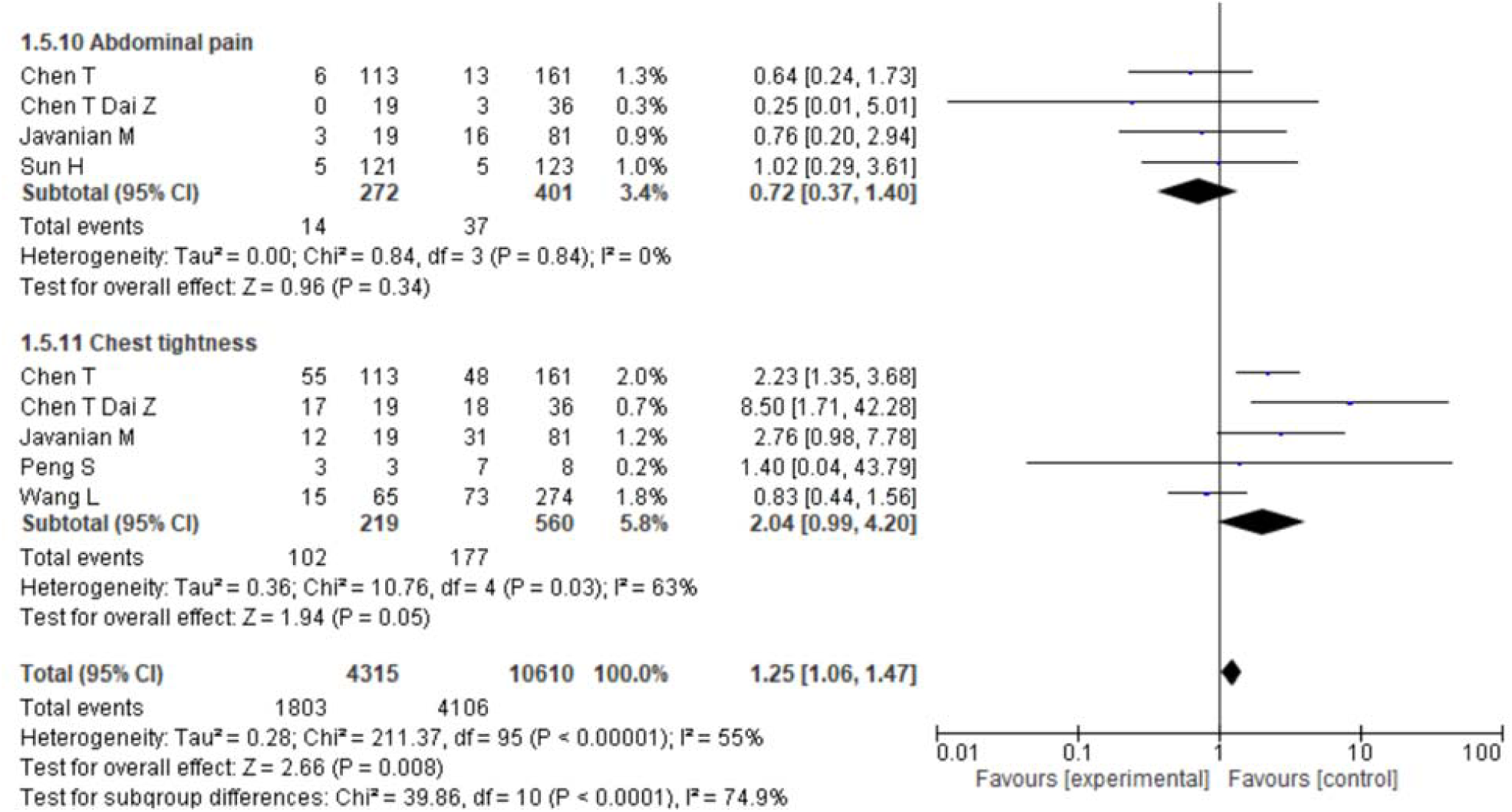
Pooled effects of clinical symptoms on the death linked with COVID-19

### 3.7 Mortality

The rate of mortality is significantly lower than the rate of survival (death vs. servival: 6.42 % vs. 93.48%) and the rate of death is 0.03-times lower than the rate of survival (OR = 0.03, 95% CI = 0.01-0.06, p <0.00001) with a significant higher heterogeneity (I^2^ = 99%, p <0.00001) (**Fig. 6**).

**Figure 6.**
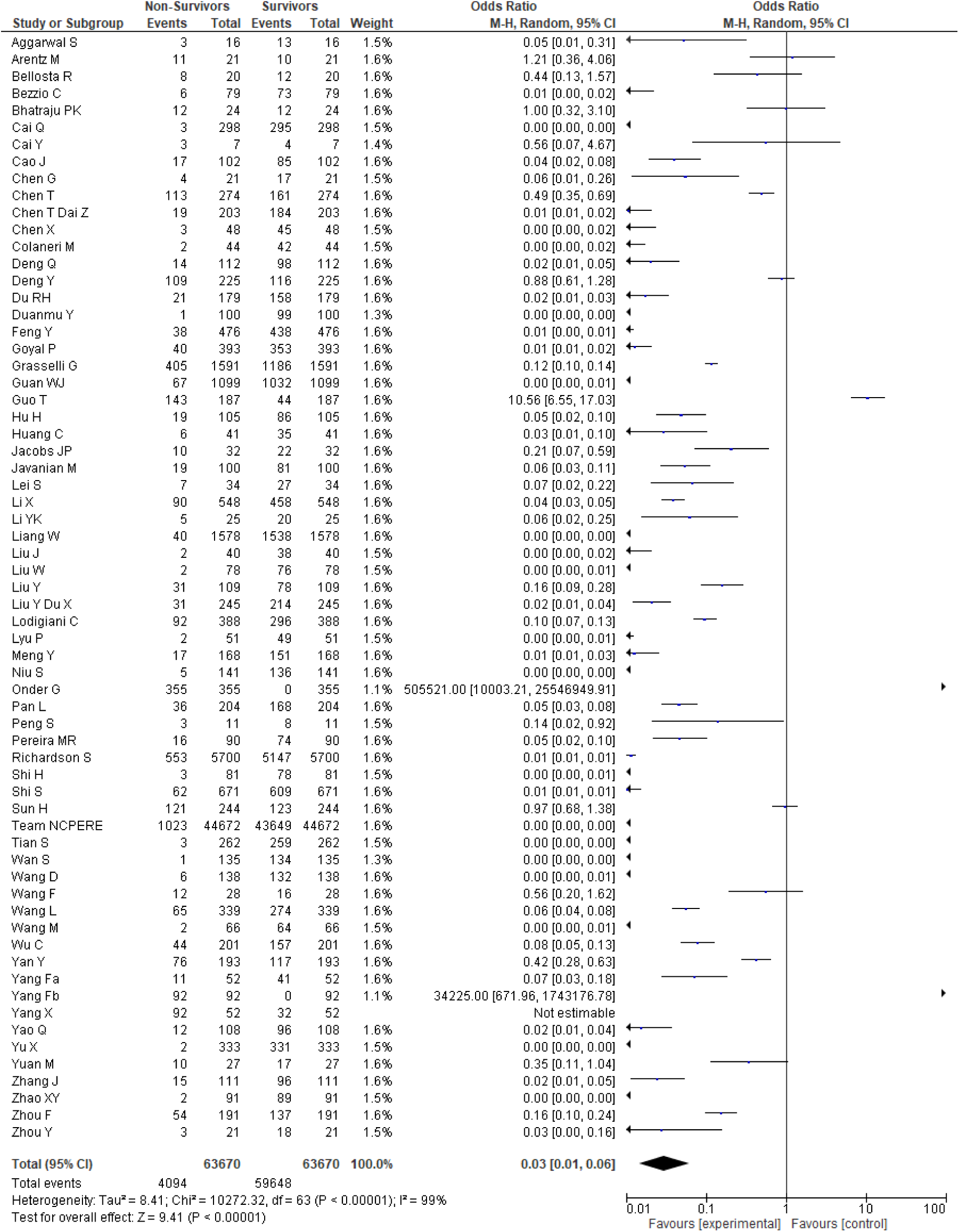
Mortality and survivors linked with COVID-19

### 3.8 Sensitivity and Publication bias

A sensitivity test was carried out by omitting the studies one by one in case of all the study parameters to assess the stability of the pooled result. No mentionable significant deviation was found in the case of all studied parameters. **Fig. 7** depicts the impact of sex on disease severity, and the odds ratio was not deviated significantly after omitting each study individually. Eagger’s test was performed to evaluate the regression asymmetry and indicated there was no notable evidence of publication bias. **Fig. 8** showed the funnel plot of malignancy as one of the comorbidity parameters.

**Figure 7.**
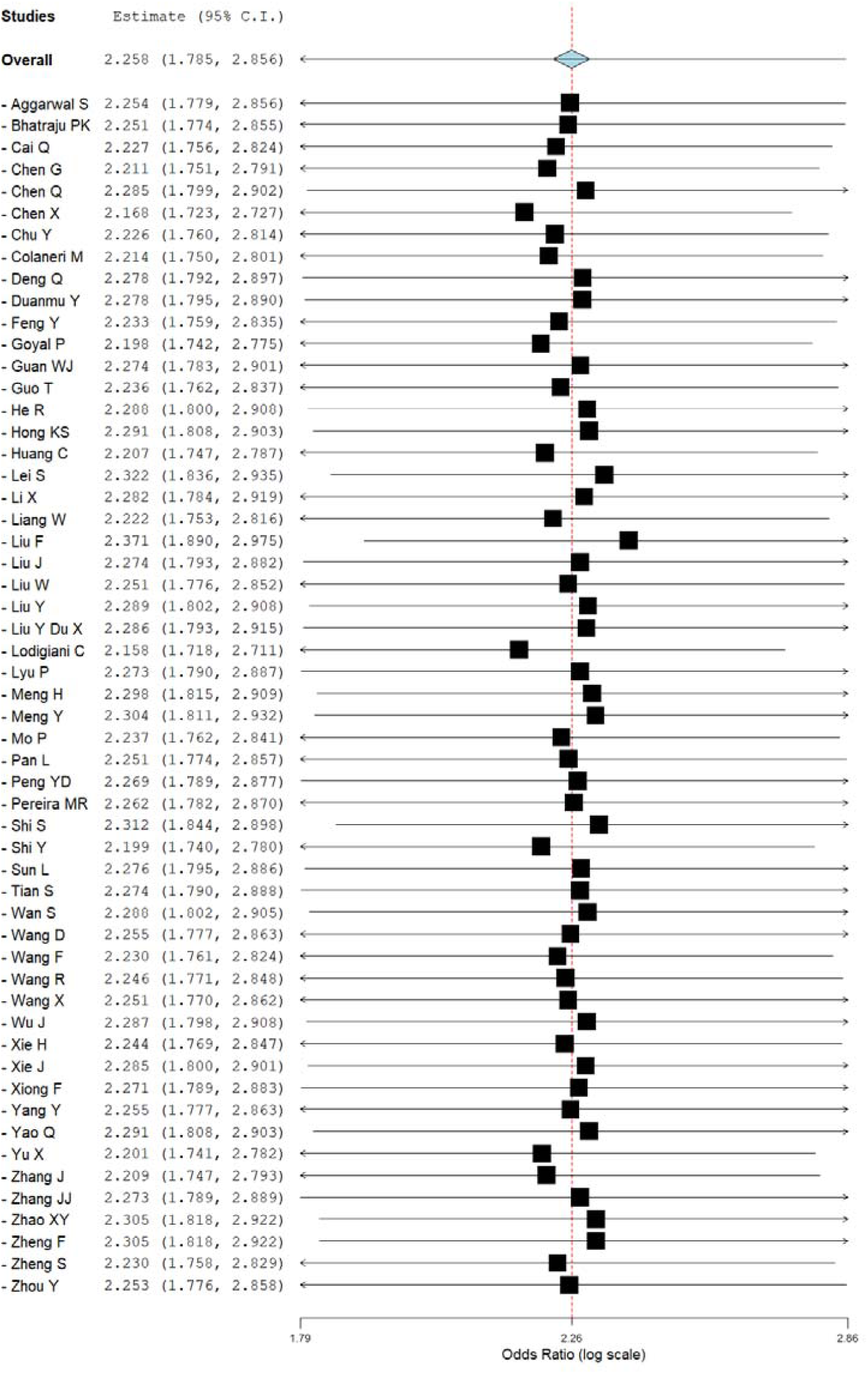
Sensitivity analysis of the effect of sex on the severity of COVID-19.

**Figure 8.**
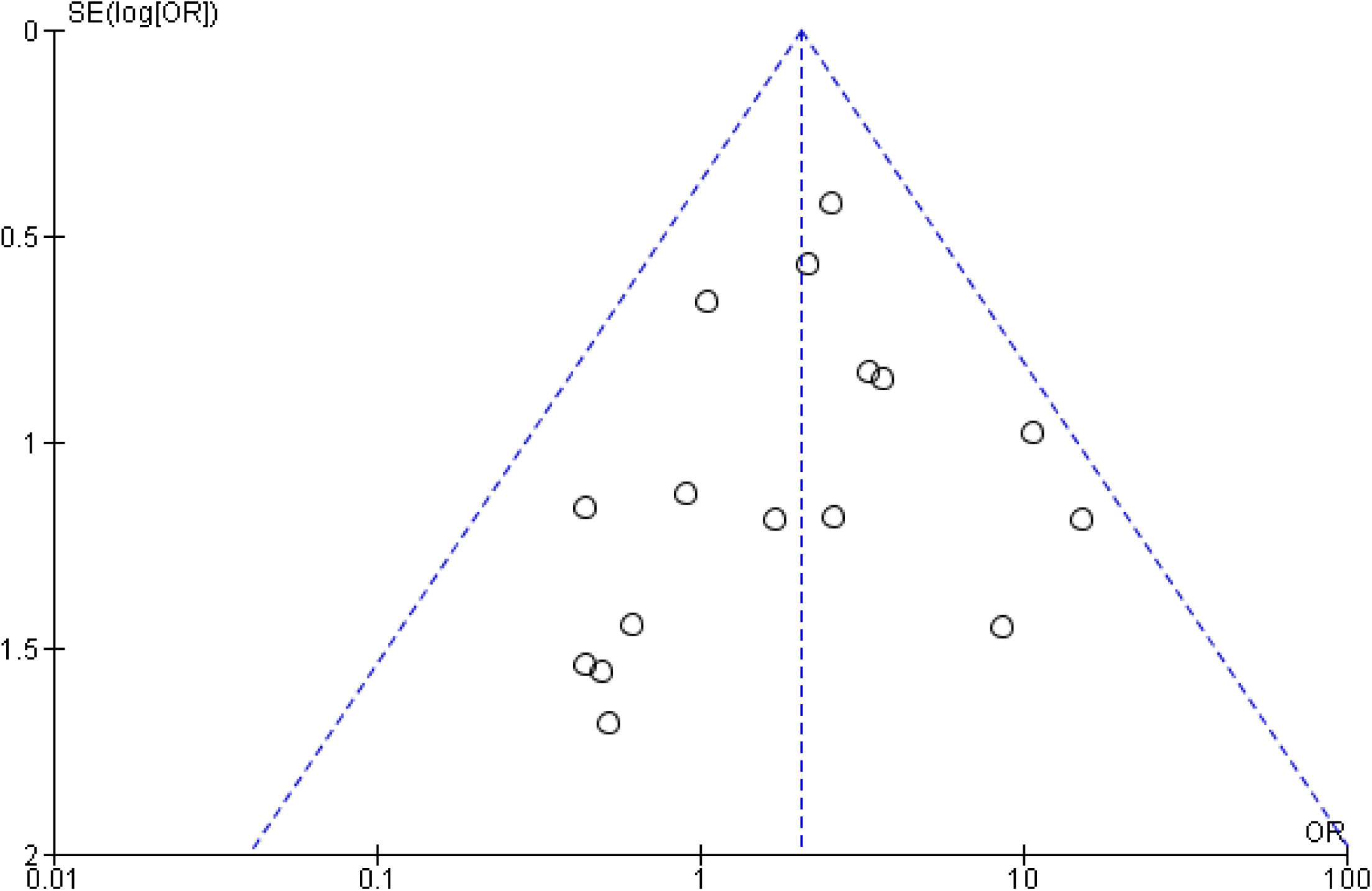
Funnel plot for the detection of publication bias

## 4. Discussion

COVID-19 is the third contagious coronavirus disease after severe acute respiratory syndrome (SARS) and the Middle East respiratory syndrome (MERS) [98]. The outbreak of COVID-19 has been declared a Public Health Emergency of International Concern by WHO. By May 17, 2020, the epidemic had spread to more than 210 countries and with the rising incidence of COVID-19 all over the world. As a newly emerging infectious disease, it is essential to identify the key clinical characteristics of COVID-19 patients to help in early detection and isolation of infected individuals, as well as minimize the spread of the disease, severity, and death rate. So, we completed a systemic meta-analysis that included 85 studies from December 2019 to May 17, 2019, to analyze prevalence and impact of age, sex, severity, comorbidity, clinical symptoms, and different outcomes with COVID-19. Our study included 5 countries, included 67,299 patients, and reflects the most recent data since the emergence of novel coronaviruses.

In our study, we have found that male patients are severely affected (OR=2.26, p < 0.00001) or died (OR=3.59, p < 0.00001) by COVID-19 rather than female and age over 50 years (≥ 50 years age vs. age<50: OR = 334.23, 95% CI =139.25-802.20, p < 0.00001) has a significantly higher risk of death. Female death or severities are lower, possibly due to the presence and protection of sex hormones and X chromosome, which play an important role in innate and adaptive immunity [99]. On the other hand, men associated with a healthy lifestyle. Several studies found that males are more likely to be infected than females during MERS-CoV and SARS-CoV pandemic [100,101]. Elderly people, severe and higher frequency of comorbidities patients are more susceptible to SARS-CoV-2 [102]. In our meta-analysis, we have found that patients having comorbidities are more prone to disease progression and death by COVID-19 comorbidities: (for at least one comorbidity carriers: death vs. survival: OR = 3.46, 95% CI =2.56-4.67, p < 0.00001). We also found that, hypertension (OR= 3.16, p < 0.00001), cardiovascular diseases (OR=4.67, p < 0.00001), diabetes (OR- 2.45, p < 0.00001), cerebrovascular disease (OR=5.84, p < 0.00001), respiratory disease (OR=2.68, p < 0.00001), liver disease (OR=2.81, p = 0.008), kidney disease (OR=5.62, p < 0.00001), malignancy (OR=2.16, p = 0.0004) were most prevalent between death groups compared to the survival group and increased the rate of death significantly compared to survival, whereas and GI disorder was not associated with the COVID related death (OR=1.43, p = 0.49). After performing the overall analysis of all comorbidities, we found that comorbidities increased the death 3.29 times compared to overall survival (OR=3.29, p<0.00001). Comorbidities and their susceptibility conditions may decrease immunity, impair macrophage and lymphocyte function, and linked to the pathogenesis of COVID-19 [103]. Heart disease patients with SARS-CoV-2 may develop a severe condition. Diabetes increases metabolic disorder and induces inflammatory infection [104]. Respiratory disorders like chronic obstructive pulmonary disease patients have lower resistance to the virus and are prone to developing ARDS and this is consistent with our findings. Hypertension and diabetes mellitus are consistent with the prevalence in China that were 21.1% and 9.7% in COVID-19 patients, respectively [103].

Furthermore, a study analyzed that diabetes, smoking, and heart disease were also significantly associated with MERS-CoV illness [105]. In this meta-analysis, we also observed that clinical manifestations-fever cough, myalgia, diarrhea, and abdominal pain were common both in death and survival patients but had significant clinical characteristics difference in fatigue (OR=1.31, p = 0.02), dyspnea (OR=4.57, p < 0.00001), and chest tightness (OR= 2.04, p = 0.05) in both groups. Fatigue and dyspnea associated significantly with an increased risk of death compared to survival. Sputum production, headache, and nausea/vomiting were higher in the death group compared with the survival group. Several clinical researchers found that the common clinical manifestations of COVID-19 patients are fever, cough, headache, fatigue, myalgia, nausea, diarrhea, and sputum and others [106, 107] that are also common in our studies. The rate of mortality found in this study also significantly lower than the survival (death vs. survival: 6.42 % vs. 93.48%, OR = 0.03, 95% CI = 0. 01-0.06, p <0.00001).

Previously, two coronavirus outbreak occurred, which are associated with the more critical disease: the SARS coronavirus in 2002-2003 and the Middle East respiratory syndrome coronavirus that have almost similar characteristics of patients with COVID-19, but there have no studies on the relation of severity and mortality with comorbidity. Similarly, there is no meta-analysis published on the severity of gender and mortality with gender, age, comorbidity, and symptoms for COVID-19 infected patients. Therefore, this is a novel systematic review and meta-analysis. Due to the lack of evidence, we present this meta-analysis by consolidating multiple retrospective studies that help to make proper and effective decisions in the future about COVID-19 patients to decrease the severity and mortality rate. It is by far the first meta-analysis with a larger sample size, and the quality of the literature included in this study is intense; the analysis is rigorous, and the conclusions drawn by the study are highly credible. However, some limitations should be mentioned. Firstly, most of the studies included in this meta-analysis were not RCTs. Secondly, high heterogeneity statistics could be found due to the larger variations in sample size. Thirdly, reports being restricted to China and a few other countries and our aim to use this study’s results to forecast patients in general, including other countries and races. Fourthly, severity, morbidity, and follow-up of patients in different hospitals vary greatly. Without these limitations, this study analyzed the risk factors for progression to critical illness or death in COVID-19 patients to help to assess patient status and identify critical patients early. Paying close attention to these risk factors and personalized treatment regimens are needed to enhance the efficacy as well as reduce the risk of death.

## 3 Conclusion

COVID-19 pandemic has led to a major health concern globally. Our results note that males and the age of more than 50 years in both sexes are at higher risk of COVID-19 infection severity and death. Most of the comorbidities, such as hypertension, diabetes, respiratory disease, cardiovascular diseases, cerebrovascular disease, renal disease, liver disease, malignancy, are responsible for increasing the COVID-19 related death. The most common symptoms of COVID-19 death and survival patients were fever, cough, fatigue, dyspnea, myalgia, diarrhea and headache, whereas fatigue and dyspnea are associated with increased death. This meta-analysis will help to make proper decisions to identify risk factors and critical patients to give suitable treatment and management promptly.

## Data Availability

All data related to this meta-analysis has been included in the manuscript.

